# Identifying Appropriate Height Exponents and the Impact on Diagnosis and Prognosis of Left Ventricular Hypertrophy: the REMODEL Study

**DOI:** 10.1101/2024.02.29.24303583

**Authors:** Clement Lee, Ryan Chan, Desiree-Faye Toh, Michelle Kui, Vivian Lee, Jennifer A Bryant, Redha Bourbetakh, Chi-Hang Lee, Chai Ping, Thu-Thao Le, Calvin WL Chin

**Author notes:** **Corresponding Author:** Calvin Chin, MD PhD FESC FRCP Senior Consultant and Clinician Scientist, NHCS Director of Cardiovascular Magnetic Resonance, NHCS Associate Professor, Cardiovascular ACP, Duke-NUS Medical School.

## Abstract

**BACKGROUND:** Left ventricular hypertrophy (LVH) is a strong predictor of adverse outcomes. Although normalizing LV mass (LVM) to height exponents reduced variability from body size, specific recommendations for height exponents are lacking due to a scarcity of normal cohorts to define appropriate height exponents. We aimed to demonstrate the diagnostic and prognostic implications of establishing height exponents specific to sex, ethnicity and imaging modality.

**METHODS:** Non-overweight/non-obese Asian healthy volunteers (n=416) were used to establish appropriate height exponents. The impact of these height exponents was examined in a separate cohort of Asians with hypertension (n=878). All individuals underwent standardized cardiovascular magnetic resonance. Primary outcome was a composite of acute coronary syndrome, heart failure hospitalization, stroke, and cardiovascular mortality.

**RESULTS:** The height exponents for healthy females and males were 1.57 and 2.33, respectively. LVH was present in 27% of individuals with hypertension when indexed to body surface area (BSA) and 47% when indexed to sex-specific height exponents. Most individuals reclassified to LVH with height exponents were overweight or obese. There were 37 adverse events over 60 (37-73) months of follow-up. Regardless of indexing method, LVH was independently associated with increased adverse events (**height exponent hazard ratio (HR)**: 2.8 [1.25-6.29], P=0.013; **BSA HR**: 5.43 [2.49-11.8], P<0.001).

**CONCLUSIONS:** Reference ranges specific to ethnicity, sex and imaging modality are necessary to establish appropriate height exponents. Although utilising height exponents resulted in more LVH reclassification, this did not translate to a notable improvement in event prediction.

Graphical Abstract
In this cardiovascular magnetic resonance study, the appropriate height exponents were 1.57 in Asian females and 2.33 in Asian males. Normalizing to height exponents increased the diagnosis of hypertensive left ventricular hypertrophy (LVH), predominantly in those who were overweight and obese. Regardless of the method of indexing, LVH was associated with adverse primary and secondary outcomes.

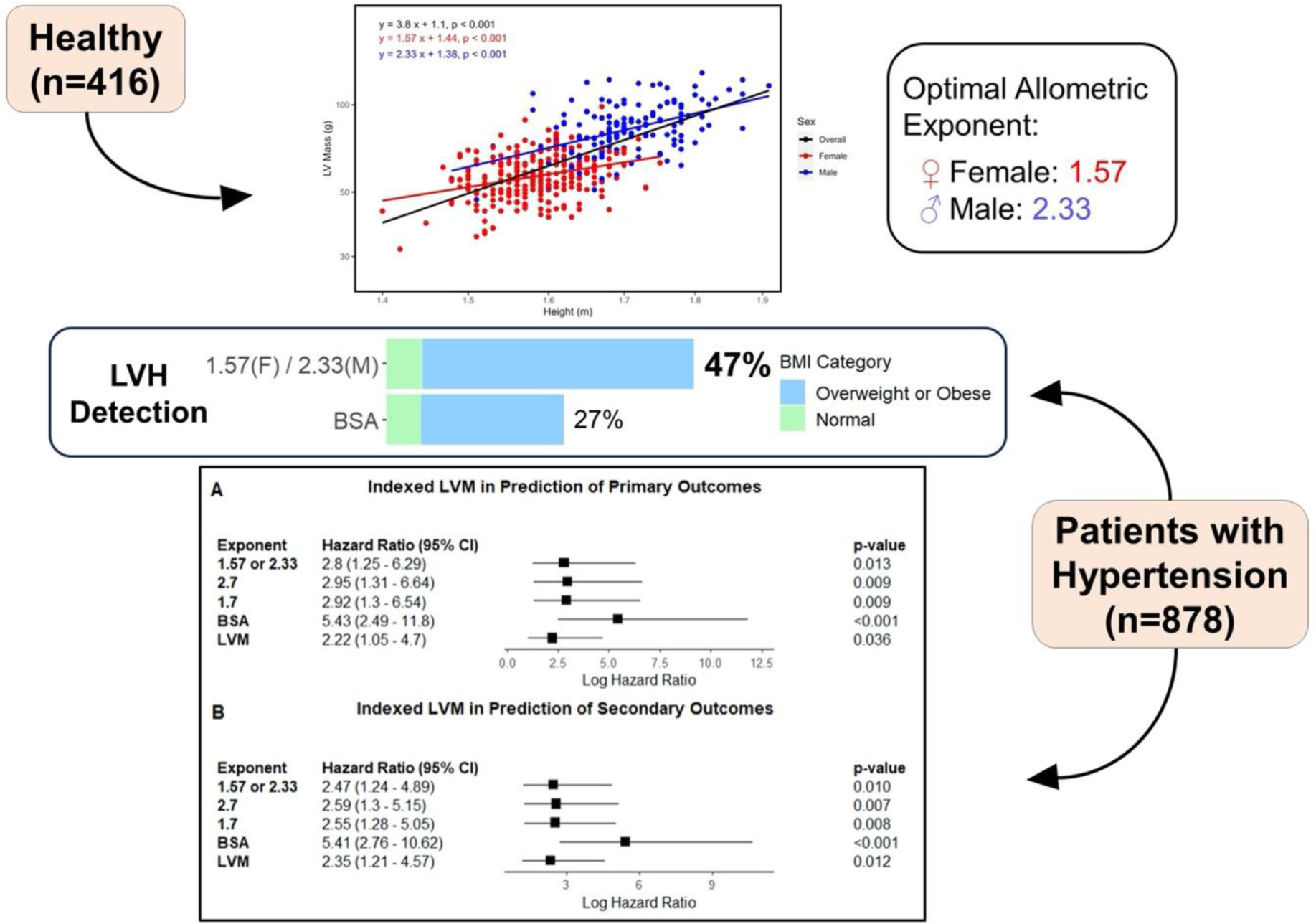

## INTRODUCTION

Left ventricular mass (LVM) is a powerful predictor of cardiovascular risk^1-3^. LVM strongly relates to body size, indicating the need for appropriate normalization. In adults, LVM is commonly indexed to body surface area (BSA), although this method has been criticised for underestimating left ventricular hypertrophy (LVH) in those who are overweight and obese^4^.

One proposed approach is normalization of LVM by height to some allometric power. For echocardiography, indexing LVM by height to the allometric power of 1.7 or 2.7 has shown the best relation to body size and events prediction^5-7^. While cardiovascular magnetic resonance (CMR) is increasingly used in assessing LVM, the existing body of research on allometric exponents in this context is focused on the Multi-Ethnic Study of Atherosclerosis (MESA) cohort^7,8^.

Although the MESA study cohort is distinguished by its ethnic diversity, the intrinsic effects of ethnicity may not fully be discerned if certain ethnic groups are under-represented. Consequently, the determination of appropriate height exponents and reference ranges contingent upon factors such as sex, ethnicity, and imaging modalities, remains inadequately elucidated. This underscores the urgent need for more precise diagnostic approaches for LVH. This aim of the study is to examine the impact of sex-specific height exponents on the diagnosis and prognosis of LVH, leveraging Asian cohorts of health and hypertension.

## METHODS

### Study Population

The study was carried out with two cohorts. The healthy cohort consisted of volunteers without cardiac symptoms, coronary artery risk factors, and clinical and family history of cardiovascular disease who were prospectively recruited from the community. Healthy volunteers who were overweight/obese (body mass index (BMI) >23.0kg/m^2^ in Asia) and had abnormal cardiac findings (regional wall motion abnormalities, impaired left ventricular function and/or cardiomyopathies) on CMR were excluded. This cohort was used to establish reference ranges of LVM indexed to sex-specific height exponents.

The impact on the diagnosis and prognosis of LVH based on LVM indexed to BSA, height exponents defined from our cohort of healthy volunteers and those published in previous studies (height^1.7^ and height^2.7^) were then tested in the REMODEL (Response of the myocardium to hypertrophic conditions in the adult population; https://www.clinicaltrials.gov; unique identifier: NCT02670031) cohort. The REMODEL is a prospective, observational cohort of asymptomatic patients with essential hypertension^9,10^. The diagnosis of hypertension was guided by contemporary recommendations at the time of study initiation: (1) physician-diagnosed essential hypertension, receiving at least one medication for blood pressure control or (2) newly diagnosed hypertension with office blood pressure ≥ 140/90 mm Hg at least two separate clinic visits. Individuals with secondary causes of hypertension, cardiovascular diseases, inherited cardiomyopathies, atrial fibrillation, and contraindications to gadolinium contrast and CMR were excluded.

Ethics approvals were obtained from the SingHealth Centralized Institutional Review Board, and all participants provided written informed consent. The study was conducted in accordance with the principles of the Declaration of Helsinki.

### Body Mass Index and Body Surface Area

In all individuals, BMI was computed as 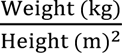. Asian BMI thresholds were used to define classifications into normal (BMI <23.0kg/m^2^), overweight (BMI 23.0 to 27.5 kg/m^2^) and obese (BMI >27.5kg/m^2^)^11^. Body surface area (BSA) was calculated using the DuBois formula^12^, as follows: BSA = weight (kg)^0.425^ x height (cm)^0.725^ x 0.007184.

### Cardiovascular Magnetic Resonance Imaging and Analysis

All study participants underwent CMR (Siemens Aera 1.5T, Siemens Healthineers, Erlangen, Germany) with the standardized imaging protocols. Relevant to this study, balanced steady state free precession (SSFP) cine images were acquired in the left ventricular (LV) long-axis 2-, 3-, and 4-chamber views (acquired voxel size, 1.6×1.3×8.0 mm^3^; 30 phases per cardiac cycle). Short axis cines extending from the atrioventricular ring to the apex were obtained to cover the entire left and right ventricles (acquired voxel size, 1.6×1.3×8.0 mm^3^; 30 phases per cardiac cycle).

Deidentified imaging data were analyzed at the National Heart Research Institute Singapore (NHRIS) CMR Core Laboratory using a dedicated software (CVI42; Circle Cardiovascular Imaging, Calgary, Canada) by individuals who were blinded to the clinical and outcome data. Cardiac volumes, function, and LV mass were analyzed according to published protocols^13^. LVH was defined according to age- and sex-specific reference ranges of LVM indexed to BSA and height exponents.

### Clinical Outcomes

Primary outcome in the REMODEL cohort was a composite of hypertension-related events, defined as acute coronary syndromes (ACS), heart failure (HF) hospitalization, stroke, and cardiovascular mortality. Secondary outcome was a composite of ACS, HF hospitalization, stroke and all-cause mortality. Recruitment started in February 2016 and participants were followed up until December 2023. Data of patients lost to follow up and did not have an event were censored at the date when patient was last known to be alive.

### Statistical Analysis

Continuous variables were assessed for normal distribution using the Shapiro–Wilk test. Data were presented in either mean ± SD or median (interquartile range), as appropriate. Analyses were performed using R (version 4.3.1, R Foundation for Statistical Computing, Vienna, Austria) and SPSS, version 24 (SPSS; IBM, Inc, Armonk, NY), assuming a 2-sided test with a 5% level of significance.

### (a) Healthy Volunteer Cohort

The relationship between LV mass and body height was modelled according to the allometric equation y = a · x^b^ where ‘y’ is LV mass in grams, ‘a’ is a constant, ‘x’ is height in meters, and ‘b’ represents the height power exponent. The ‘x’ and ‘y’ variables logarithmically transformed: ln (y) = ln (a) + b · ln (x) ^14^. Subsequently, ordinary least squares linear regression was performed to estimate the allometric scaling exponent, ‘b’.

Separate simple linear regression analyses were used to model the association between LVM and age in males and females. Reference ranges were defined using the 95% prediction interval: mean ± t_0.975, n-1_ (√(n+1)/n)·(SD). To account for the sample size, 95% confidence intervals of the (upper and lower) reference limits were also estimated. Values within these confidence intervals were considered “indeterminate abnormal/borderline normal”^15^.

### (b) REMODEL Cohort

Time-to-event analysis of primary and secondary outcomes in hypertensive patients with LVH (unindexed, indexed to BSA and height exponents) was performed using multivariable Cox proportional hazards models adjusted for diabetes, hyperlipidemia, systolic blood pressure (SBP). The model of unindexed LVM was additionally adjusted for height and weight. The assumption for proportional hazards was assessed using the log (−log[survival]) plots.

## RESULTS

The study consisted of 416 healthy volunteers (48±15 years; females, n=257) and 878 patients with hypertension (58±11 years; females, n=336; SBP 131±14mmHg) (**Table 1**).

**Table 1.** Baseline Characteristics of Healthy Volunteers and Patients with Hypertension. Abbreviations: SBP, systolic blood pressure; DBP diastolic blood pressure; LV left ventricular

|  | **Healthy**<br>**Volunteers**<br>**(n=416)** | **Hypertension**<br>**Patients**<br>**(n=878)** | **P value** |
| --- | --- | --- | --- |
| **Clinical Characteristics** |  |  |  |
| Age, years | 47.8±15.4 | 58.0±10.6 | <0.001 |
| Female, n (%) | 257 (62) | 336 (38) | <0.001 |
| Hyperlipidemia, n (%) | - | 429 (49) | - |
| Diabetes, n (%) | - | 188 (21) | - |
| 24-h SBP, mmHg | - | 131±14 | - |
| 24-h DBP, mmHg | - | 79±9 | - |
| Office SBP, mmHg | 127±19 | 143±18 | <0.001 |
| Office DBP, mmHg | 75±12 | 83±12 | <0.001 |
| Body Surface Area, m<sup>2</sup> | 1.60±0.10 | 1.80±0.20 | <0.001 |
| **Left Ventricular Mass Measures on Cardiovascular Magnetic Resonance** |  |  |  |
| LV Mass, g | 67±18 | 96±32 | <0.001 |
| LV Mass, g/m<sup>2</sup> | 42±9 | 53±14 | <0.001 |
| LV Mass, g/height<sup>1.7</sup> | 29±6 | 40±12 | <0.001 |
| LV Mass, g/height<sup>2.7</sup> | 18±4 | 24±7 | <0.001 |
| LV Mass, g/height<sup>1.57 (f)/2.33 (m)</sup> | 27±5 | 33±9 | <0.001 |

### Derivation of Height Exponents and Reference Ranges

Simple linear regression was performed after logarithmic transformation of LV mass, and height. The height exponents for healthy females and males were found to be 1.57 and 2.33, respectively (**Figure 1**). Sex-specific reference ranges in LVM, both unindexed and indexed to BSA and height exponents were established (**Table 2**). When normalized to height exponents, females had higher LVM compared to males (28±5g/h^1.57^ versus 24±4g/h^2.33^, P<0.001). Conversely, when normalized to BSA, females had lower LVM compared to males (38±6g/m^2^ versus 49±8g/m^2^, P<0.001) (**Table 2**).

**Figure 1.**
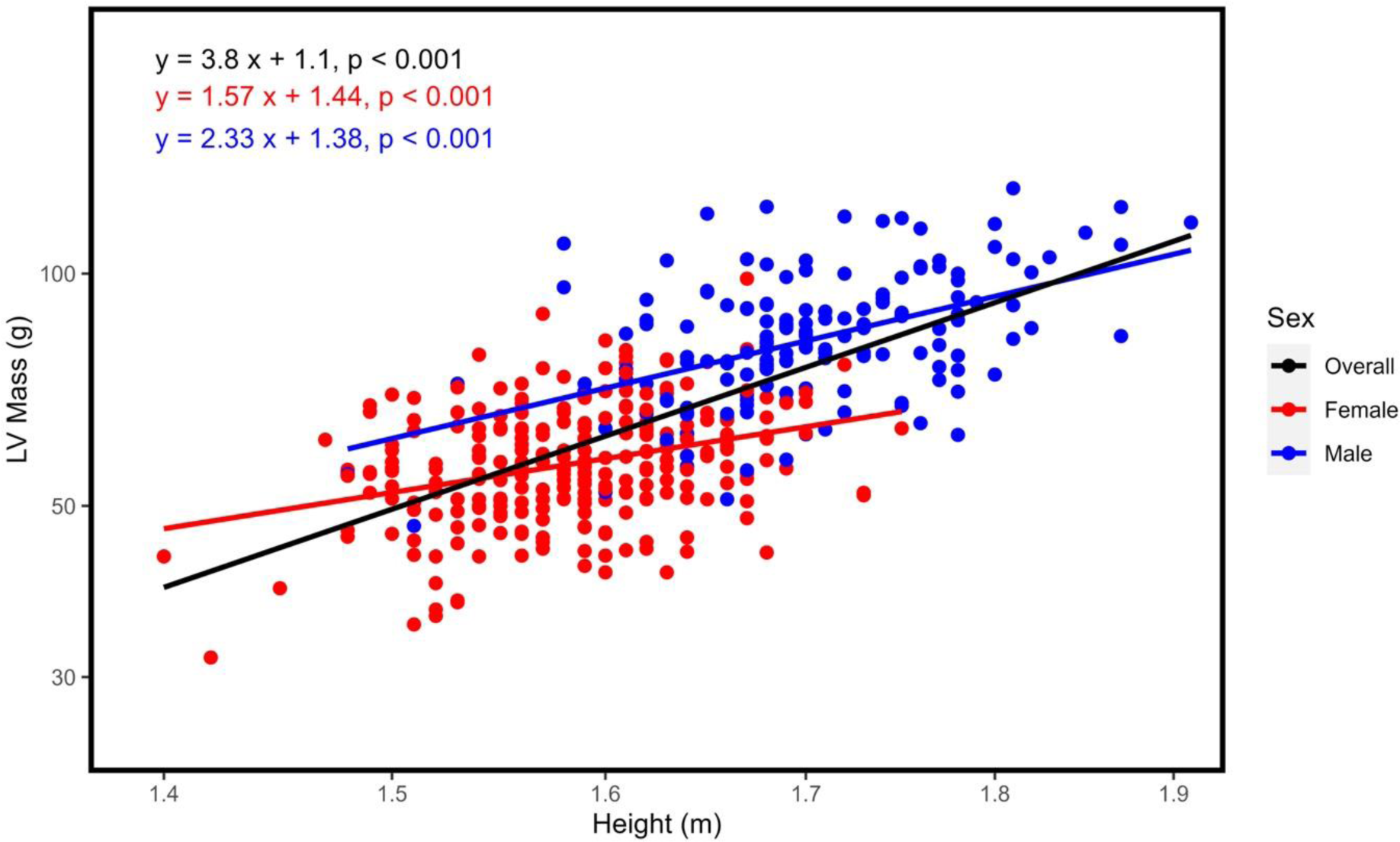
Association between Left Ventricular Mass and Height. The relationship between left ventricular (LV) mass and height can be modelled by the allometric equation y = a · x ^b^, where ‘y’ is LV mass in grams, ‘x’ is height in meters and ‘b’ represents the height exponent. Logarithmic transformations of LV mass and height were performed. The height exponent for the overall healthy cohort was 3.38. The sex-specific height exponent was 1.57 for females and 2.33 for males.

**Table 2.**
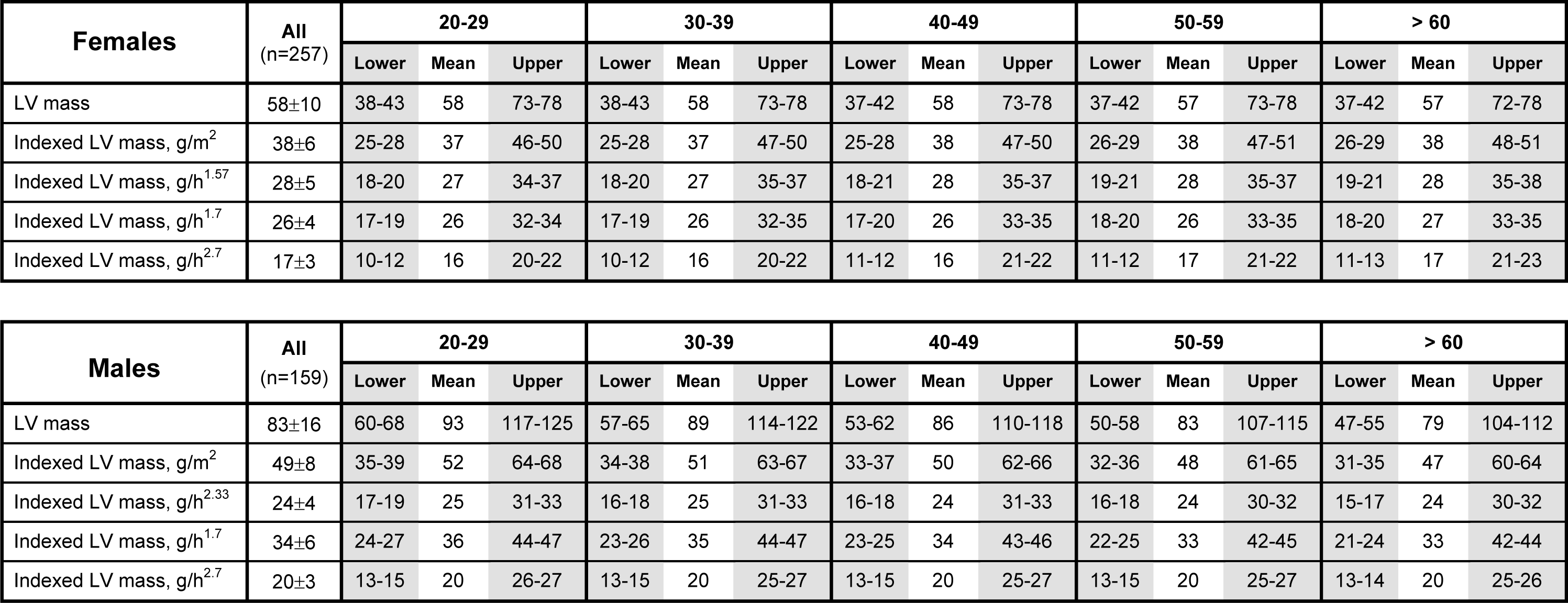
Sex-specific Reference Ranges in Left Ventricular Mass, Unindexed and Indexed to Body Surface Area and Height Exponents. Values given in the upper and lower confidence intervals of the reference limits (grey columns) are considered “indeterminate abnormal/borderline normal”. The lower bound of the upper reference limit was used as the diagnostic cut-off for left ventricular hypertrophy in this study. Abbreviations: LV left ventricular.

| Females | All<br>(n=257) | 20-29 |  |  | 30-39 |  |  | 40-49 |  |  | 50-59 |  |  | > 60 |  |  |
| --- | --- | --- | --- | --- | --- | --- | --- | --- | --- | --- | --- | --- | --- | --- | --- | --- |
|  |  | Lower | Mean | Upper | Lower | Mean | Upper | Lower | Mean | Upper | Lower | Mean | Upper | Lower | Mean | Upper |
| LV mass | 58±10 | 38-43 | 58 | 73-78 | 38-43 | 58 | 73-78 | 37-42 | 58 | 73-78 | 37-42 | 57 | 73-78 | 37-42 | 57 | 72-78 |
| Indexed LV mass, g/m<sup>2</sup> | 38±6 | 25-28 | 37 | 46-50 | 25-28 | 37 | 47-50 | 25-28 | 38 | 47-50 | 26-29 | 38 | 47-51 | 26-29 | 38 | 48-51 |
| Indexed LV mass, g/h<sup>1.57</sup> | 28±5 | 18-20 | 27 | 34-37 | 18-20 | 27 | 35-37 | 18-21 | 28 | 35-37 | 19-21 | 28 | 35-37 | 19-21 | 28 | 35-38 |
| Indexed LV mass, g/h<sup>1.7</sup> | 26±4 | 17-19 | 26 | 32-34 | 17-19 | 26 | 32-35 | 17-20 | 26 | 33-35 | 18-20 | 26 | 33-35 | 18-20 | 27 | 33-35 |
| Indexed LV mass, g/h<sup>2.7</sup> | 17±3 | 10-12 | 16 | 20-22 | 10-12 | 16 | 20-22 | 11-12 | 16 | 21-22 | 11-12 | 17 | 21-22 | 11-13 | 17 | 21-23 |

| Males | All<br>(n=159) | 20-29 |  |  | 30-39 |  |  | 40-49 |  |  | 50-59 |  |  | > 60 |  |  |
| --- | --- | --- | --- | --- | --- | --- | --- | --- | --- | --- | --- | --- | --- | --- | --- | --- |
|  |  | Lower | Mean | Upper | Lower | Mean | Upper | Lower | Mean | Upper | Lower | Mean | Upper | Lower | Mean | Upper |
| LV mass | 83±16 | 60-68 | 93 | 117-125 | 57-65 | 89 | 114-122 | 53-62 | 86 | 110-118 | 50-58 | 83 | 107-115 | 47-55 | 79 | 104-112 |
| Indexed LV mass, g/m<sup>2</sup> | 49±8 | 35-39 | 52 | 64-68 | 34-38 | 51 | 63-67 | 33-37 | 50 | 62-66 | 32-36 | 48 | 61-65 | 31-35 | 47 | 60-64 |
| Indexed LV mass, g/h<sup>2.33</sup> | 24±4 | 17-19 | 25 | 31-33 | 16-18 | 25 | 31-33 | 16-18 | 24 | 31-33 | 16-18 | 24 | 30-32 | 15-17 | 24 | 30-32 |
| Indexed LV mass, g/h<sup>1.7</sup> | 34±6 | 24-27 | 36 | 44-47 | 23-26 | 35 | 44-47 | 23-25 | 34 | 43-46 | 22-25 | 33 | 42-45 | 21-24 | 33 | 42-44 |
| Indexed LV mass, g/h<sup>2.7</sup> | 20±3 | 13-15 | 20 | 26-27 | 13-15 | 20 | 25-27 | 13-15 | 20 | 25-27 | 13-15 | 20 | 25-27 | 13-14 | 20 | 25-26 |

### Impact on the Diagnosis of Left Ventricular Hypertrophy in Individuals with Hypertension

In the REMODEL cohort, LVH was present in 236 (27%) and 409 (47%) individuals when LVM was indexed to BSA and sex-specific height exponents, respectively.

Across BMI categories, classification of LVH was concordant in 80% of individuals (n=701) between LVM indexed to BSA and sex-specific height exponents. LVM indexed to height exponents reclassified 175 individuals to having LVH, with more reclassifications across higher BMI categories: normal BMI, n=4 (2.3%); overweight, n=58 (33.1%); obese, n=113 (64.6%). Conversely, only two individuals (both with normal BMI) were reclassified to having LVH when LVM was indexed to BSA (**Figure 2**).

**Figure 2.**
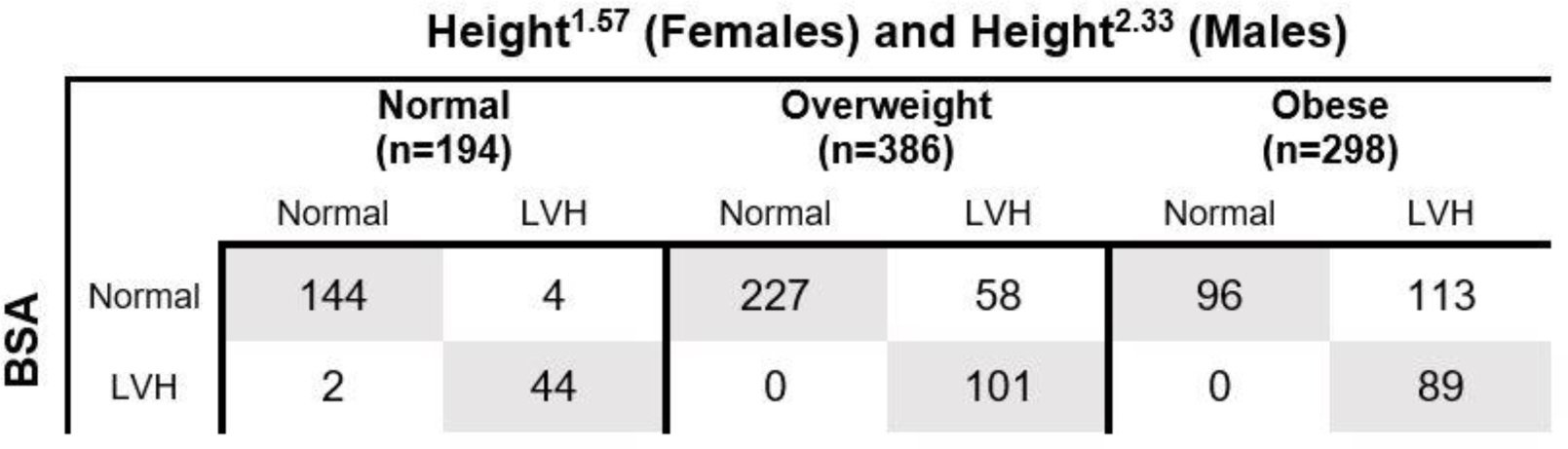
Left Ventricular Hypertrophy Classification Based on Myocardial Mass Indexed to Body Surface Area and Height Exponents. Left ventricular hypertrophy (LVH) diagnosed based on LV mass indexed to body surface area (BSA), or height exponents (1.57 in females and 2.33 in males) was concordant in 80% of patients (grey cells). A total of 175 patients were reclassified as having LVH when height exponents were used, with the majority being in the overweight (n=58; 33.1%) and obese (n=113; 64.6%) categories.

### Prognostic Value of Left Ventricular Hypertrophy Using Sex-Specific Height Exponents

There were 37 primary and 47 secondary outcomes over 60 (37-73) months of follow-up: ACS, n=12; HF hospitalization, n=8, stroke, n=12; cardiovascular deaths, n=5, and non-cardiovascular deaths, n=10.

We examined the prognosis of LVH diagnosed with unindexed LVM, indexed to BSA and height exponents (including height^1.7^ and height^2.7^). Regardless of the method of diagnosis, LVH was independently associated with increased primary and secondary outcomes. LVH diagnosed based on LV mass indexed to BSA was associated with the highest numerical HR, but its 95% confidence intervals overlapped with those of other LVH measures (**Figure 3**).

**Figure 3.**
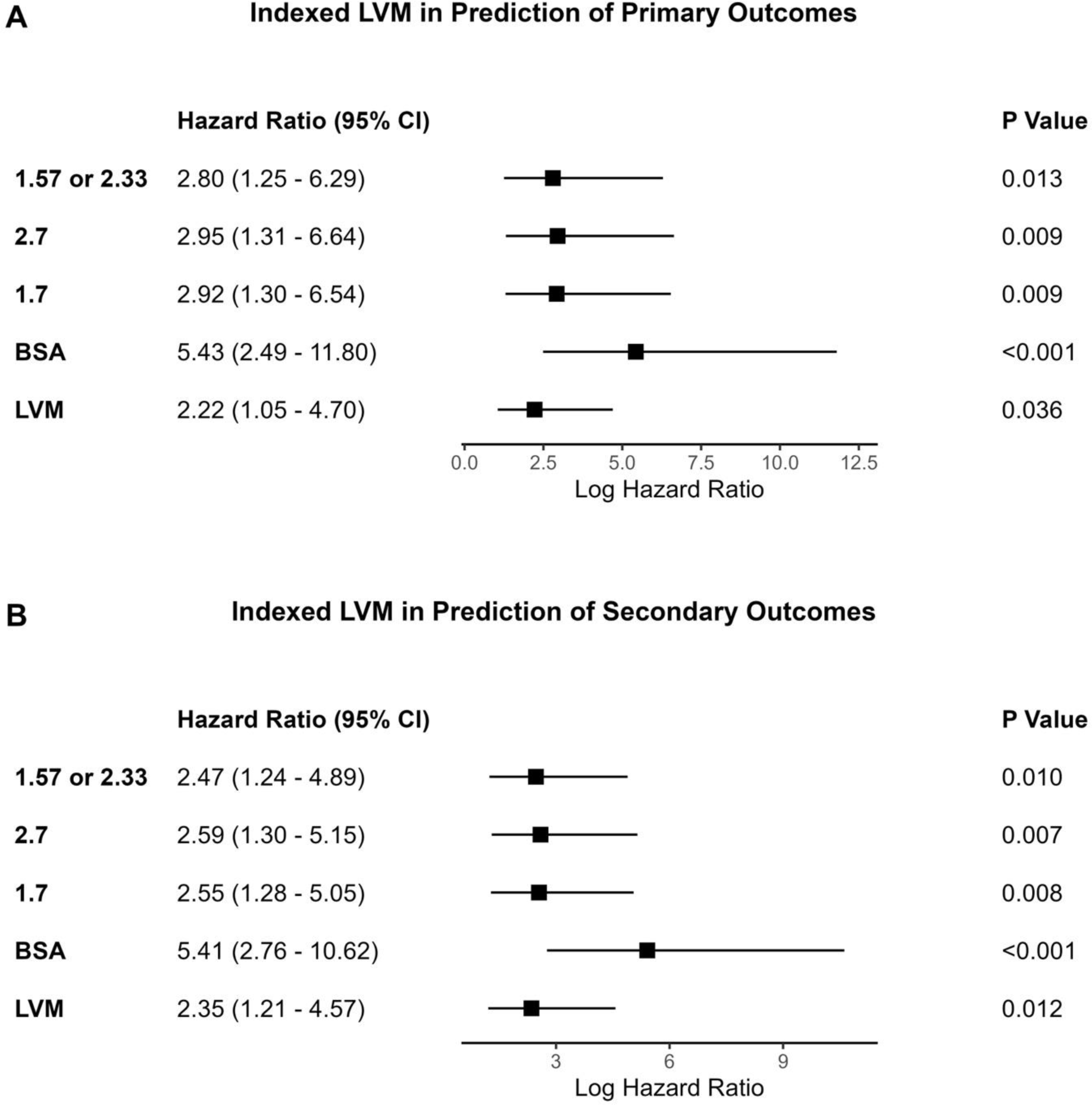
Impact of Left Ventricular Hypertrophy Measures on Outcomes. Cox models of left ventricular hypertrophy (LVH) measures were adjusted for 24-hour systolic blood pressure, diabetes, and hyperlipidemia. LVH diagnosed with unindexed left ventricular mass (LVM) was additionally adjusted for height and weight. All LVH measures were significantly and independently associated with worse primary (**Panel A**) and secondary (**Panel B**) outcomes.

The number of primary outcomes was similar in those with LVH regardless of the diagnostic method used. Of the 37 primary outcomes, 25 occurred in those with LVH based on unindexed LVM, 27 in those with LVH diagnosed using LVM indexed to BSA and 29 in individuals with LVH diagnosed using height exponents. Furthermore, the distribution of primary outcomes related to LVH across BMI categories were comparable regardless of the approach of indexing (**Table 3**). Similar findings were observed with secondary outcomes (**Table 3**).

**Table 3.** Primary and Secondary Outcomes in Individuals with Left Ventricular Hypertrophy Stratified by Body Mass Index. The number of primary and secondary outcomes associated with LVH were similar regardless of the approach of indexing and across strata of body mass index. Abbreviations: BMI, body mass index; BSA body surface area; LVM unindexed left ventricular mass

|  |  | Primary Outcome |  |  |  |  |  |
| --- | --- | --- | --- | --- | --- | --- | --- |
|  |  | LVM | BSA | Height<sup>1.57 (f)</sup><br>/Height<sup>2.33 (m)</sup> | Height<sup>1.7</sup> | Height<sup>2.7</sup> | P Value |
| BMI category | Normal, n (%) | 7 (28) | 8 (30) | 8 (28) | 8 (28) | 8 (28) | >0.99 |
|  | Overweight, n (%) | 11 (44) | 12 (44) | 13 (45) | 13 (45) | 13 (45) |  |
|  | Obese, n (%) | 7 (28) | 7 (26) | 8 (28) | 8 (28) | 8 (28) |  |

|  |  | Secondary Outcome |  |  |  |  |  |
| --- | --- | --- | --- | --- | --- | --- | --- |
|  |  | LVM | BSA | Height<sup>1.57 (f)</sup><br>/Height<sup>2.33 (m)</sup> | Height<sup>1.7</sup> | Height<sup>2.7</sup> | P Value |
| BMI category | Normal, n (%) | 7 (23) | 8 (24) | 8 (23) | 8 (23) | 8 (23) | >0.99 |
|  | Overweight, n (%) | 14 (45) | 15 (45) | 16 (46) | 16 (46) | 16 (46) |  |
|  | Obese, n (%) | 10 (32) | 10 (30) | 11 (31) | 11 (31) | 11 (31) |  |

## DISCUSSION

Despite the potential merit of allometric scaling, current guidance on appropriate scaling coefficients is lacking. Furthermore, there is a paucity of studies to critically assess the influence of ethnicity on allometric scaling and outcomes. In this CMR study, height exponents in Asians differed between sexes (2.33 in males and 1.57 in females), contrasting with the single height exponent of 1.7 or 2.7 reported in previous studies. Utilizing these exponents in individuals with hypertension resulted in increased reclassification of individuals with hypertensive LVH, particularly among those who were overweight and obese. However, this alternative approach of indexing LVM did not translate to significant improvement in event prediction (**Graphical Abstract**).

### Allometric Scaling Improves the Diagnosis of LVH

Ideally, LVM should be normalized to lean/fat-free muscle mass. A clinically practical and convenient surrogate of fat-free mass is body height, and allometric height-exponent indexing of LVM potentially overcomes the limitation of underestimating LVH in overweight or obese individuals. Indeed, the study demonstrated nearly a two-fold increased in LVH prevalence when height-exponent indexing of LVM was used. These results were consistent with previous studies which reported how LVM indexed to height, height^2.13^, or height^2.7^ diagnosed significantly more LVH than when indexed to BSA^4,16-19^. Also consistent with previous studies, a large proportion of the patients who were reclassified to having LVH based on height-exponent indexing were overweight or obese^4,8,20^.

### Prognostic Impact of Different LVH Measures

In echocardiographic studies, indexing LVM to height^1.7^ or height^2.7^ have shown the best relationship to body size and event prediction^5-7^. CMR is widely recognized as an important imaging modality in diagnosing LVH because of its superior accuracy and precision in assessing LVM^21^. The two CMR studies used the MESA cohort to examine the importance of LVM indexed to height exponents and reported a higher prevalence of LVH for measures that did not account for weight, but there were no important differences in prognostic value among the indices studied^7,8^.

Compared to these contemporary CMR studies, the current study had a larger cohort of a homogeneous ethnicity and with a wider age range (20 to 88 years old versus 45 to 84 in the MESA cohort). We demonstrated that LVH diagnosed based on height-exponent and BSA indexing were similar in predicting adverse outcomes. Although the HR of LVH indexed to BSA was numerically higher, the confidence interval overlapped with those indexed to height exponents. This was in agreement with studies that reported similar cardiovascular events and mortality risk regardless of either BSA or height-exponent indexing were used^7,16,17,19^.

### Clinical Implications

In non-obese individuals, the approach of normalization will likely not have significant impact on the diagnosis of LVH. In contrast, in populations with a high prevalence of obesity, indexing with BSA underestimates the effects of obesity and reduces the prevalence of LVH. Our findings endorse the importance of normalizing LVM by height exponents in such a scenario. For this application, it is crucial the appropriate reference ranges specific to the ethnicity, sex and imaging modality be established.

Our study reported the diagnosis of LVH, whether based on either BSA or height-exponent indexing, was concordant in 80% of patients with hypertension. Moreover, the distribution of adverse outcomes related to LVH across BMI categories were similar regardless of the indexing approach. Whilst height-exponent indexing maximizes the population-attributable risk, this consideration must also be balanced against the lack of a significant improvement in event prediction.

### Study Limitations

We acknowledge the study was limited by a relatively small number of events. However, we believe this limitation is unlikely to significantly compromise the validity of the study. The observed findings are consistent with existing knowledge and the confidence intervals surrounding most point estimates were relatively narrow. While our investigation demonstrated that indexing LVM to height exponents reduced variability attributed to body size and sex, the allometric exponents established in this study warrant further validation and comparison with other ethnicities. Such efforts will enhance the generalizability and robustness of the findings across different demographic profiles.

## PERSPECTIVES

Normal reference ranges specific to the ethnicity, sex and imaging modality are necessary to establish appropriate height exponents. Our study reported that Asian males and females have unique height exponents, differing from those observed in earlier studies. Although the adoption of height exponents resulted in increased reclassification of individuals with LVH, this did not translate to a notable improvement in event prediction.

## Data Availability

Data will be available upon reasonable request to the corresponding author

## ABBREVIATIONS

ACS: Acute Coronary Syndrome
BMI: Body Mass Index
BSA: Body Surface Area
CMR: Cardiovascular Magnetic Resonance
HF: Heart Failure
LVH: Left Ventricular Hypertrophy
LVM: Left Ventricular Mass
SBP: Systolic Blood Pressure

## ACKNOWLEDGEMENTS

We thank the radiographers at the Department of Cardiovascular Magnetic Resonance, National Heart Centre Singapore, for performing the scans in the study.

